# Establishing laboratory reference ranges for adults and children in Kilifi, Kenya

**DOI:** 10.1101/2024.10.08.24315083

**Authors:** Louise O Downs, Benedict Orindi, Mainga Hamaluba, Philip Bejon, Lynette Isabella Ochola-Oyier, Caroline Ngetsa

**Author notes:** **Corresponding author:** Louise Downs.

## Abstract

Accurate laboratory reference ranges (RR) are essential for diagnosis and management of patients in routine clinical care and clinical trials. RRs vary between geographical location due to differences in population demographics and blood analysis equipment, so locally derived RRs are essential. Here we establish adult and paediatric RRs for a rural population in Kilifi, Kenya using clinical trial data from KEMRI-Wellcome Trust Research Programme (KWTRP).

Data from healthy, non-pregnant participants from six clinical trials conducted between 2016 and 2020 were used. Coulter ACT 5 Diff and Ilab Aries were used for haematological and biochemical analysis respectively. Quality control was undertaken daily prior to sample analysis. Derived RRs were compared with RRs from other African countries and further afield. All analyses were performed using R version 3.6.1 (Reference Intervals package).

2338 adults and 2054 children were included, 52% of adults and 51% of children were male, median adult age was 32.5 years. Haemoglobin range was lower in women compared to men (9.5–14.2g/dL and 11.5–16.6g/dL respectively), platelet upper limit of normal (ULN) was higher in women compared to men (397 × 10^3^/μL vs 358 × 10^3^/ μL). Biochemistry values were higher in men (ALT ULN 57 U/L in men and 35 U/L in women, creatinine ULN 113umol/L in men and 91umol/L in women). Paediatric RRs showed differences in multiple parameters depending on the age of the child.

In both adults and children, many parameters in 2023 Kilifi RRs differed from those in other countries. There was however little difference between 2023 and 2017 Kilifi paediatric RRs.

This study provides RRs for adults and children in Kilifi, and the most extensive RRs available for much of East and Southern Africa. We show the need for locally derived reference ranges, highlighting differences between sex, age and geographical location.

## Introduction

The reference range (RR) for a biological parameter is the range of test values expected for a designated population where at least 95% of the individuals are presumed to be healthy (1). RRs are powerful tools to aid decision-making in clinical practice and defining RRs enables identification of any future observation from the population which falls outside of this range, and hence may require further investigation (1). It also allows monitoring of disease progression and treatment response.

In clinical trials, RRs are used to determine eligibility for trial participation and for grading of toxicity or adverse events, usually combined with a clinical assessment of significance. International guidelines from the Clinical and Laboratory Standards Institute (CLSI) advise determining local ranges where possible (2). It has previously been shown there are significant differences in certain parameters both between different African populations and between the Global North and African populations (3–5) and this can significantly influence the proportion of people thought to have abnormal results in Africa, potentially leading to unnecessary investigations or exclusion from clinical trials. This may result from genetic differences (for instance a greater pool of marginating neutrophils in benign “ethnic” neutropenia) or from different exposures (for instance lower platelet counts due to recent malaria infection). This leads to challenges in recruitment, prolonging research time and increasing financial costs. Consequently, many countries have derived region specific RRs (4,6–8).

The Kenya Medical Research Institute (KEMRI)-Wellcome Trust Research Programme (KWTRP) in Kilifi, coastal Kenya, is a large research institute conducting multiple clinical studies annually, investigating both communicable and non-communicable diseases. Numbers of clinical trials undertaken in East Africa have increased significantly over the last 10 years (9) and there is a need to ensure locally applicable blood RRs are established for correct result interpretation. Normal RRs vary depending on both laboratory factors such as the machines used and types of blood collection bottles, and population factors such as age, sex, diet, environment and ethnicity. The population of Kilifi County is predominantly rural and of lower socio-economic status than other Kenyan counties which is likely to influence RRs and highlights the need for geographically specific ranges. We previously undertook paediatric reference range determination in 2017 but only included children up to 17 months and did not determine reference ranges for adults (10). The numbers of biochemical parameters included for children was also limited, looking at only creatinine and alanine aminotransferase (ALT).

Here we collate haematological and biochemical parameters from healthy volunteers from six clinical trials at KWTRP aiming to: i) develop adult and paediatric RRs for the Kilifi population, ii) expand the available paediatric reference ranges up to 18 years, iii) increase the number of biochemical parameters available locally for reference, and iv) compare these results with laboratory results from other locations in Kenya and internationally. This work provides some of the most extensive RRs for biochemical parameters in Africa, and to our knowledge, no other ranges have subdivided paediatric ranges as extensively as is done here.

## Methods

### Study design, setting and data sources

This was a secondary analysis of four clinical trials conducted at KWTRP in Kilifi between 2016–2020 to derive adult RRs (11–14), and a further two studies were included for paediatric RRs (15,16). The full details of the data collection protocols and findings have previously been extensively described (11–16). Briefly, each trial randomly selected participants from the Kilifi Health and Demographic Surveillance System (KHDSS) study area and clinically assessed them for chronic disease. All participants were also screened for Hepatitis B, Hepatitis C, malaria and HIV before trial enrolment and excluded if found positive. Pregnant or breastfeeding women were excluded from trials and women of childbearing age were asked to take effective contraception throughout trial duration. Following consent and trial enrolment, baseline blood samples were collected from participants and used to derive these RRs. Different blood parameters were tested for each study based on the study aims, so not all parameters were available for all participants of all studies. These newly defined RRs are referred to as ‘Kilifi 2023’ throughout this manuscript, recognising that the studies took place prior to 2023.

### Laboratory analysis

Haematology and biochemistry samples were analysed in the KWTRP clinical trials laboratory (CTL). The CTL was accredited in 2006 by Qualogy LTD and has since maintained good clinical and laboratory practice (GCLP) compliance.

Haematological complete blood count was performed using the Beckman Coulter ACT 5 Diff analyser from blood collected in ethylenediamine tetra acetic acid (EDTA). The parameters analysed included red blood cells (RBC), haemoglobin (Hb, g/dL), haematocrit (HTC, %), mean cell volume (MCV, fL), platelets (PLT, 10^3^/ μL), white blood cells (WBC, 10^3^/μL), neutrophils (10^3^/μL), lymphocytes (10^3^/μL), monocytes (10^3^/μL), basophils (10^3^/μL) and eosinophils (10^3^/μL).

Biochemistry tests were performed in blood collected in serum separating tubes using the Ilab Aries analyser for alanine aminotransferase (ALT, U/L), aspartate aminotransferase (AST), creatinine, albumin, blood glucose, gamma glutamyl transferase (GGT), inorganic phosphate, urea, sodium (Na), potassium (k) magnesium (mg).

### Quality control

Three levels of quality control samples; high, low and normal were run daily before sample analysis for both haematology and biochemistry tests to ensure the analysers were producing reliable results. The CTL is also enrolled on two different external quality assurance schemes; United Kingdom’s National External Quality Assurance Services (UKNEQAS) and the Royal College of Pathologists of Australasia (RCPA) which ensures results are accurate, reliable and comparable. The equipment was on service contract and routinely serviced by qualified service providers as per the recommended maintenance schedule.

### Populations used for comparisons

To review how these 2023 RRs compared with those of other countries both within the African continent and further afield, we identified studies that analysed a similar combination of reference markers over similar age ranges. The parameters compared were those most widely available, and some of those most commonly shown to have significant geographical variation. For adults, the RRs from Kisumu also included adults between 18-35 years, excluded those with identifiable chronic illness, and included a population which was predominantly rural (17). The CLSI Southern Africa RRs included populations from multiple African countries, with a similar median population age to these 2023 Kilifi RRs at 28 years. Again those with identifiable chronic illness were excluded, and there was a higher proportion of urban living than in Kilifi (18). For paediatric comparisons, it was challenging to identify RRs within Africa with a similar age split and parameter measurement to these 2023 Kilifi RRs. Firstly, we compared with the 2017 Kilifi RRs which were derived from a similar rural paediatric population across Kilifi and were analysed using the same haematological analyser but different biochemical analyser. This enabled us to review whether these new 2023 ranges differed from those previously derived from a similar population. We then reviewed international RRs from both Canada and the United Kingdom primarily to highlight differences and to exemplify why having location specific RRs is essential. These populations were both urban and children with known chronic illness were excluded.

### Statistical methods

All analyses were performed using R version 3.6.1 (R Core Team, 2019) making use of the reference Intervals package (Daniel, 2014). We eliminated outlying observations for each parameter, defined as being more than 1.5 times the interquartile range above the third quartile or below the first quartile. Given the skewed distribution for most measures, we used a non-parametric approach to estimate the 95% RRs following the CLSI C28-A3 guidelines, partitioned by sex. For each upper and lower bound, a 90% confidence interval was constructed using bootstrapping method.

### Ethical considerations

All parent studies received ethical approval from KEMRI-Scientific Ethics Review Unit (SERU) and relevant ethical bodies for collaborating institutes, and regulatory approval from the Pharmacy and Poisons Board of Kenya. SERU also approved the plans to reuse these data for the purposes described in this manuscript. All participants in parent studies consented to reuse of their data without further consent. Consent was sought from study principal investigators and/or sponsors if necessary for reuse of the data. Data were stripped of any patient-identifiers before analysis and/or sharing (19), so authors did not have access to information that could identify individual participants. Data was accessed on 21^st^ March 2023 for the analysis included in this manuscript.

## Results

### Study populations

In these 2023 Kilifi RRs, we analysed data from 2338 adults and 2054 children. 1220/2338 (52%) adults and 1046/2054 (51%) of children were male. Adult ages ranged from 18 – 98 years although the median age for each study tended to be young (27 years in the CHMI study, 28 years in the Vac 072 study and 29 years in the S4V01 study). Only the ShinDa 2 study had a median adult age of over 30 years at 46 years. For paediatric RRs, ages ranged from 0.9 months – 17 years. Numbers included from each study are shown in **table 1**. Numbers available for each parameter varied since not all studies measured all parameters.

**Table 1:** Details of each clinical study at KEMRI-Wellcome Trust Research Programme used to determine healthy adult and paediatric haematological and biochemical reference ranges for use in clinical trials in Kilifi, Kenya.

| Study Name | Group Included | Number included in these ranges | Median age in years (range) | Male (%) |
| --- | --- | --- | --- | --- |
| CHMI: Controlled Human Malaria Infection (CHMI) to assess human immunity to *P. falciparum* using sporozoites (11). | Adults | 373 | 27 (18–45) | 289 (77.5) |
| Vac 073: A Phase 1b, open-label, age de-escalation, dose-escalation study to evaluate the safety and immunogenicity of different doses of a candidate malaria vaccine (42). | Adults | 220 | 28 (19–43) | 164 (74.6) |
|  | Children | 131 | 1 (0–5) | 62 (47.3) |
| S4V01: Safety and immunogenicity of a Shigella-tetavalent bioconjugate vaccine (12). | Adults | 35 | 29 (19–48) | 22 (62.9) |
|  | Children | 68 | 3 (0–5) | 36 (52.9) |
| ShinDa 2: Investigating the role of malaria in elevating blood pressure and pulse wave velocity in Kenyan children and adults (13). | Adults | 1710 | 46<br>(18–98) | 745<br>(43.6) |
|  | Children | 522 | 16<br>(11–17) | 299<br>(57.3) |
| **Study Name (paediatric population only)** |  |  | **Median age in months (range)** |  |
| FPCV: The Effect of Fractional Doses of Pneumococcal Conjugate Vaccines on Immunogenicity and Carriage in Kenyan Infants (43) | Children | 39 | 1.4<br>(1.2– 1.8) | 15<br>(38.5) |
| RTS,S malaria vaccine study | Children | 1294 | 8.4<br>(0.9 – 26.7) | 634<br>(49.0) |
| Total | **Adults** | **2338** |  | 1220<br>(52) |
|  | **Children** | **2054** |  | 1046<br>(51) |

For adults, haematology parameters had a sample size ranging from 398 (eosinophils) to 1103 (Mean Corpuscular Volume) individuals for females (**table 2a**) and 700 (basophils) to 1232 (Mean Corpuscular Volume) individuals for males (**table 2b**), meeting the recommended CLSI sample size of 120 (2). Most biochemical parameters also met these criteria, although some parameters less routinely measured (i.e. albumin, bilirubin and AST) were below this recommended sample size, and this should be borne in mind during interpretation (**tables 3a and 3b**).

**Table 2a:**
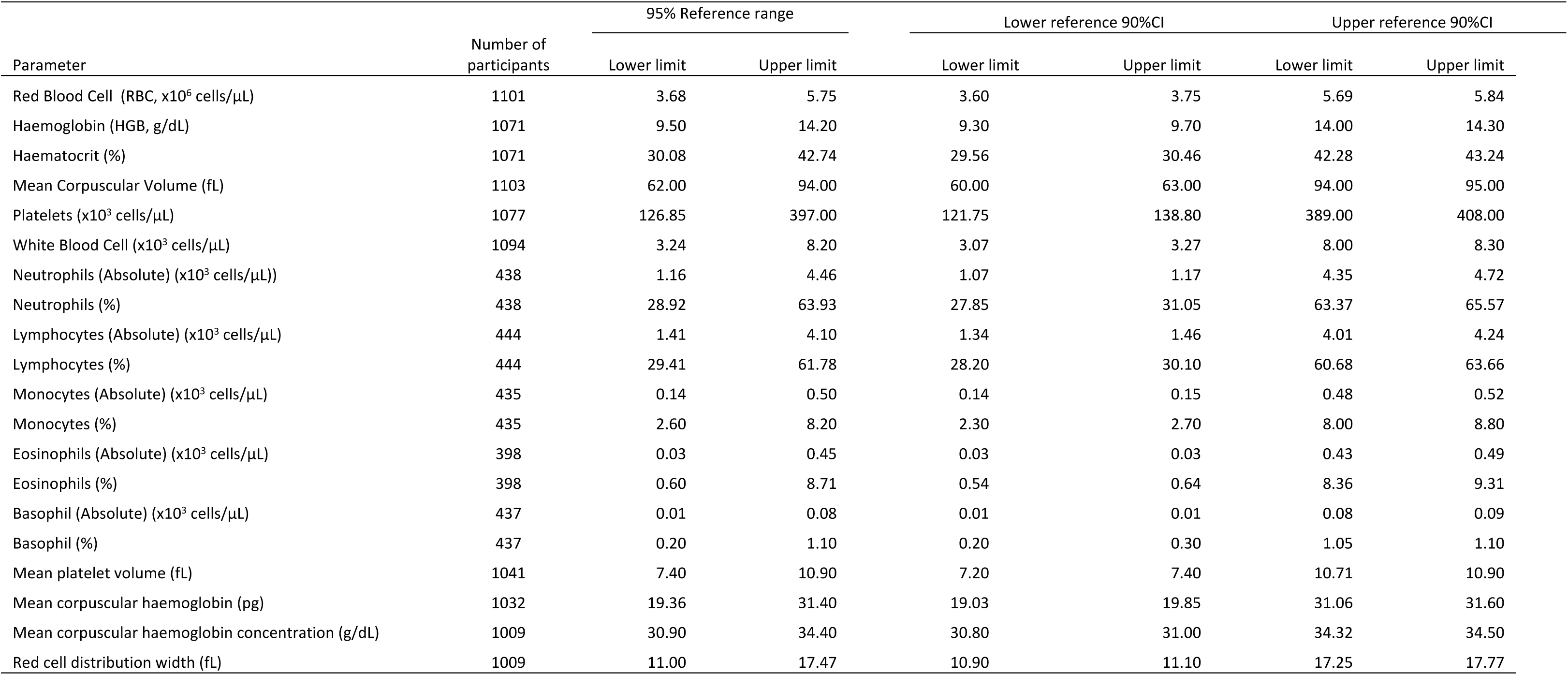
Reference ranges and associated 90% confidence intervals for **haematological** measures for healthy **adult females** in Kilifi, Kenya, derived using data from clinical trials at KEMRI-Wellcome Trust Research Programme.

**Table 2b:**
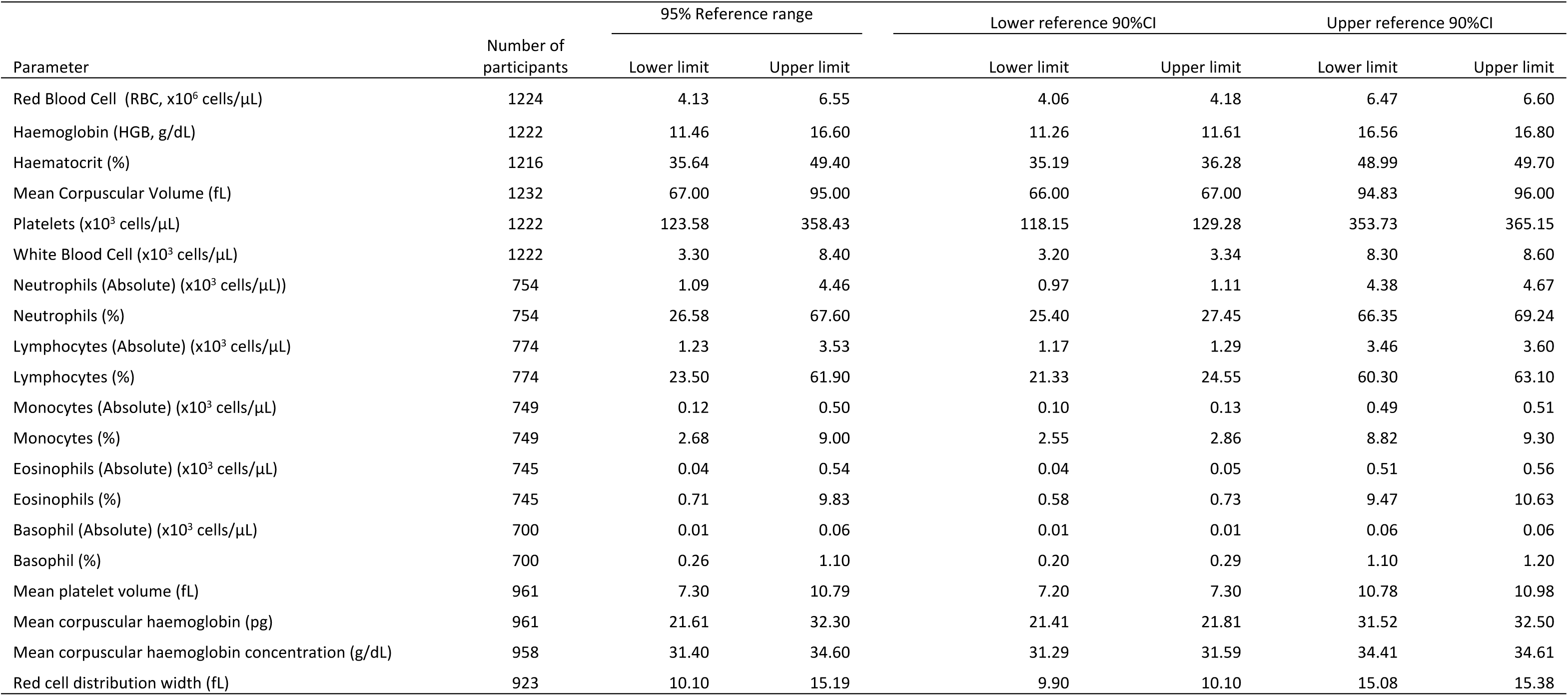
Reference ranges and associated 90% confidence intervals for **haematological** measures for healthy **adult males** in Kilifi, Kenya, derived using data from clinical trials at KEMRI-Wellcome Trust Research Programme.

**Table 3a:**
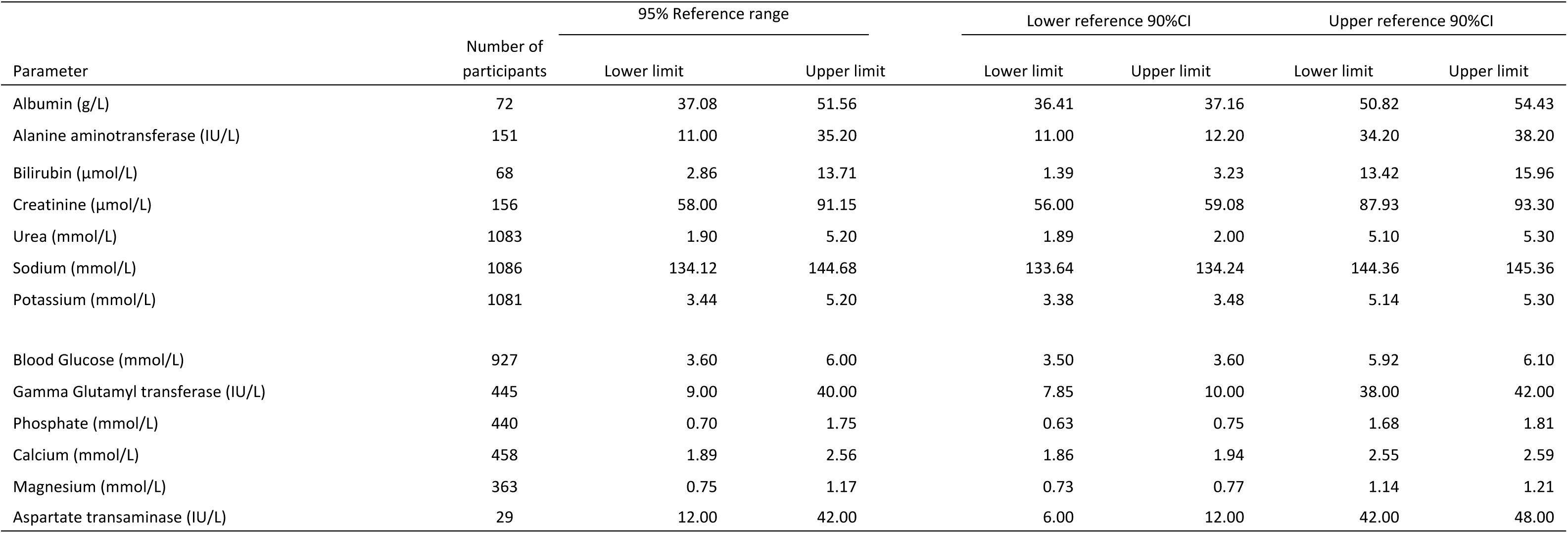
Reference ranges and associated 90% confidence intervals for **biochemistry** measures for healthy **adult females** in Kilifi, Kenya, derived using data from clinical trials at KEMRI-Wellcome Trust Research Programme.

**Table 3b:** Reference ranges and associated 90% confidence intervals for **biochemical** measures for healthy **adult males** in Kilifi, Kenya, derived using data from clinical trials at KEMRI-Wellcome Trust Research Programme.

| Parameter | Number of participants | 95% Reference range |  | Lower reference 90%CI |  | Upper reference 90%CI |  |
| --- | --- | --- | --- | --- | --- | --- | --- |
|  |  | Lower limit | Upper limit | Lower limit | Upper limit | Lower limit | Upper limit |
| Albumin (g/L) | 271 | 38.62 | 52.84 | 38.28 | 39.97 | 51.74 | 53.66 |
| Alanine aminotransferase (IU/L) | 471 | 13.80 | 57.00 | 13.60 | 14.60 | 55.00 | 59.00 |
| Bilirubin (µmol/L) | 258 | 4.54 | 24.37 | 4.08 | 5.03 | 23.32 | 25.88 |
| Creatinine (µmol/L) | 496 | 67.43 | 115.00 | 64.43 | 69.85 | 111.43 | 117.00 |
| Urea (mmol/L) | 1160 | 1.80 | 5.50 | 1.71 | 1.81 | 5.40 | 5.69 |
| Sodium (mmol/L) | 1149 | 133.00 | 143.53 | 132.70 | 133.00 | 143.05 | 144.03 |
| Potassium (mmol/L) | 1178 | 3.5 | 5.35575 | 3.47 | 3.56 | 5.28 | 5.41 |
| Glucose (mmol/L) | 706 | 3.50 | 5.90 | 3.40 | 3.60 | 5.80 | 6.00 |
| Gamma glutamyl transferase (IU/L) | 348 | 13.00 | 58.00 | 11.28 | 14.00 | 55.00 | 61.73 |
| Phosphate (mmol/L) | 351 | 0.62 | 1.64 | 0.56 | 0.68 | 1.61 | 1.69 |
| Calcium (mmol/L) | 361 | 1.91 | 2.55 | 1.90 | 1.97 | 2.52 | 2.60 |
| Magnesium (mmol/L) | 300 | 0.60 | 1.16 | 0.56 | 0.65 | 1.16 | 1.19 |
| Aspartate transaminase (IU/L) | 62 | 20.58 | 47.43 | 20.15 | 21.15 | 46.85 | 50.43 |

### Estimated Kilifi 2023 reference ranges

Within Kilifi, when comparing results by sex, there was some variation. Amongst haematological ranges, of those who had their haemoglobin and platelets measured there was an equal split between men and women (47% women, 53% men). The haemoglobin RR was however considerably lower in women (9.5–14.2g/dL) compared with men (11.5–16.6g/dL; **Tables 2a and 2b**, **Fig 1),** and platelets were higher in women compared to men (upper limit of normal (ULN) 397 × 10^3^/μL vs 358 × 10^3^/ μL (**Fig 1**)). Within biochemistry, ALT and creatinine were both lower in women (ALT ULN = 35 U/L, creatinine ULN = 91umol/L) than in men (ALT ULN = 57 U/L, creatinine ULN = 113umol/L), (**Tables 3a and 3b**, **Fig 1**). It should be noted however that of those who had their ALT and creatinine measured, considerably more were men than women (76% men vs 24% women).

**Fig 1:**
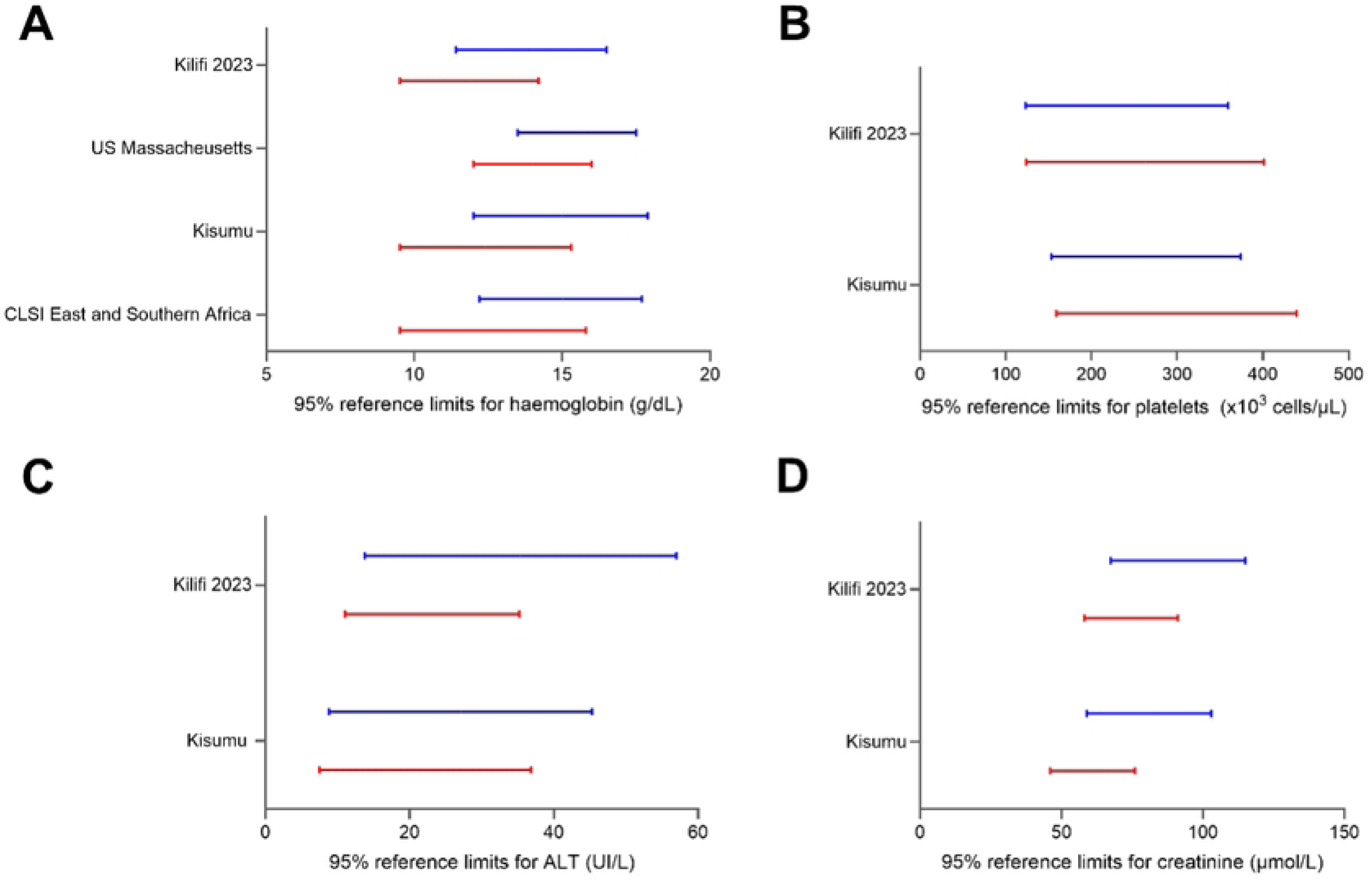
Kilifi 2023 95% adult reference ranges for common parameters (A) haemoglobin, (B) platelets, (C) ALT and (D) creatinine compared with ranges from other sites in Kenya and internationally by sex. Males are shown in blue, and females are shown in red.

Different paediatric age groups within Kilifi also showed differences in multiple parameters depending on the age of the child **(tables 4a-d and 5a-d).** Most strikingly were platelets whose ULN decreased from 752 × 10^3^/μL in those aged 1–5 months to 451 × 10^3^/ μL in those aged 60–215 months and WBCs for which the ULN changed from 14.9 × 10^3^/ μL at 1–5 months down to 8.5 × 10^3^/μL at 60–215 months.

**Table 4a:**
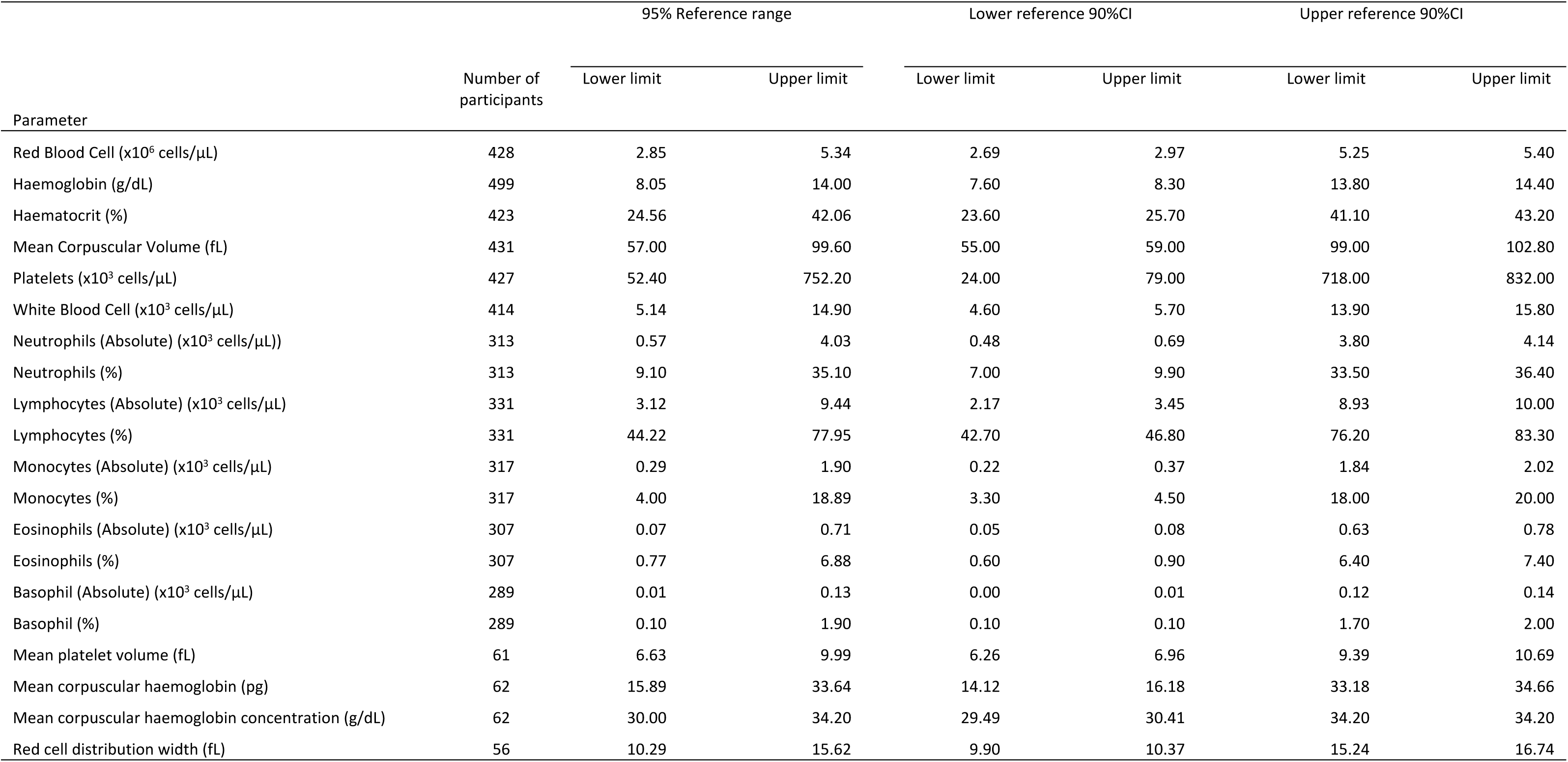
Reference ranges and associated 90% confidence intervals for **haematological** measures for healthy **children aged 1-5 months** in Kilifi, Kenya, derived using data from clinical trials at KEMRI-Wellcome Trust Research Programme.

**Table 4b:**
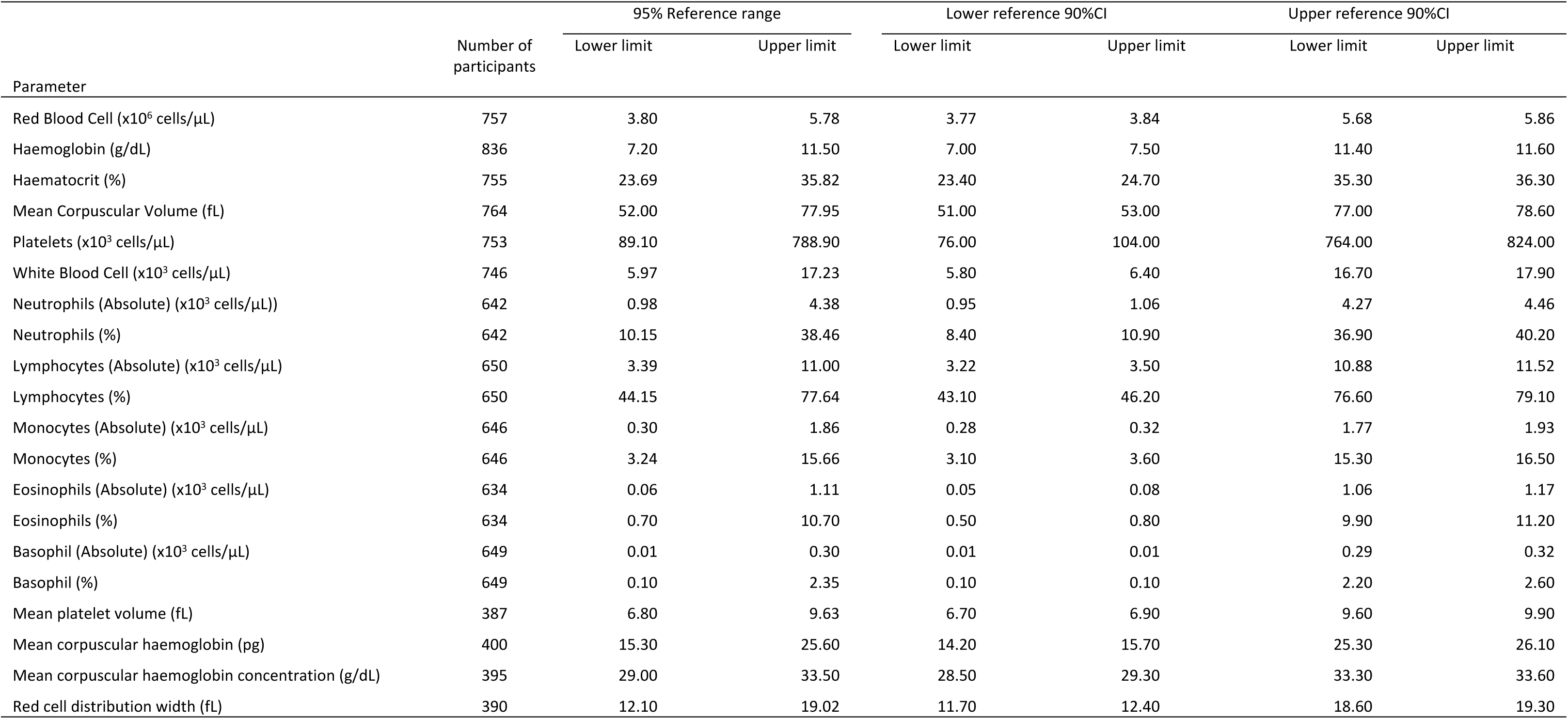
Reference ranges and associated 90% confidence intervals for **haematological** measures for healthy **children aged 6-11 months** in Kilifi, Kenya, derived using data from clinical trials at KEMRI-Wellcome Trust Research Programme.

**Table 4c:**
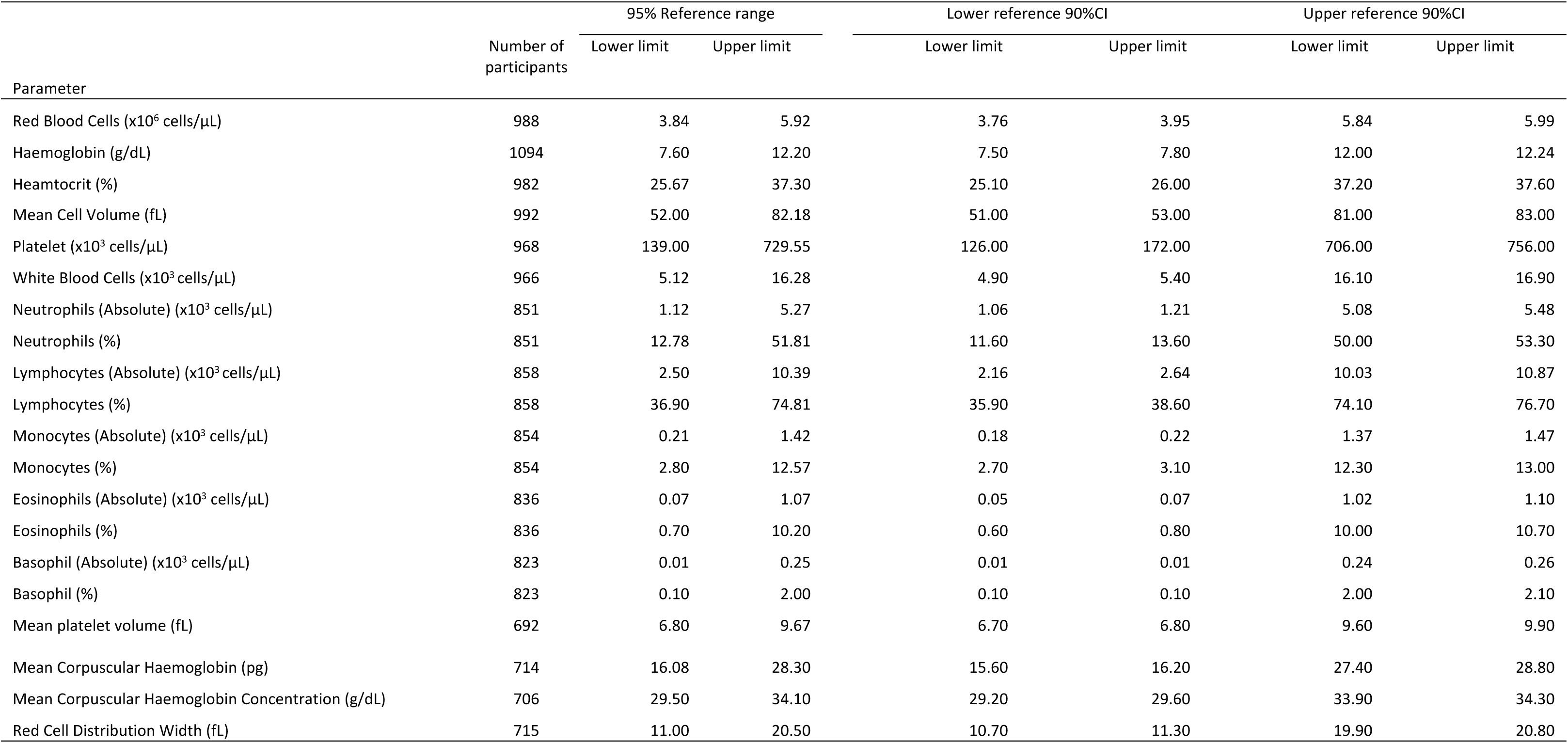
Reference ranges and associated 90% confidence intervals for **haematological** measures for healthy **children aged 12-59 months** in Kilifi, Kenya, derived using data from clinical trials at KEMRI-Wellcome Trust Research Programme.

**Table 4d:**
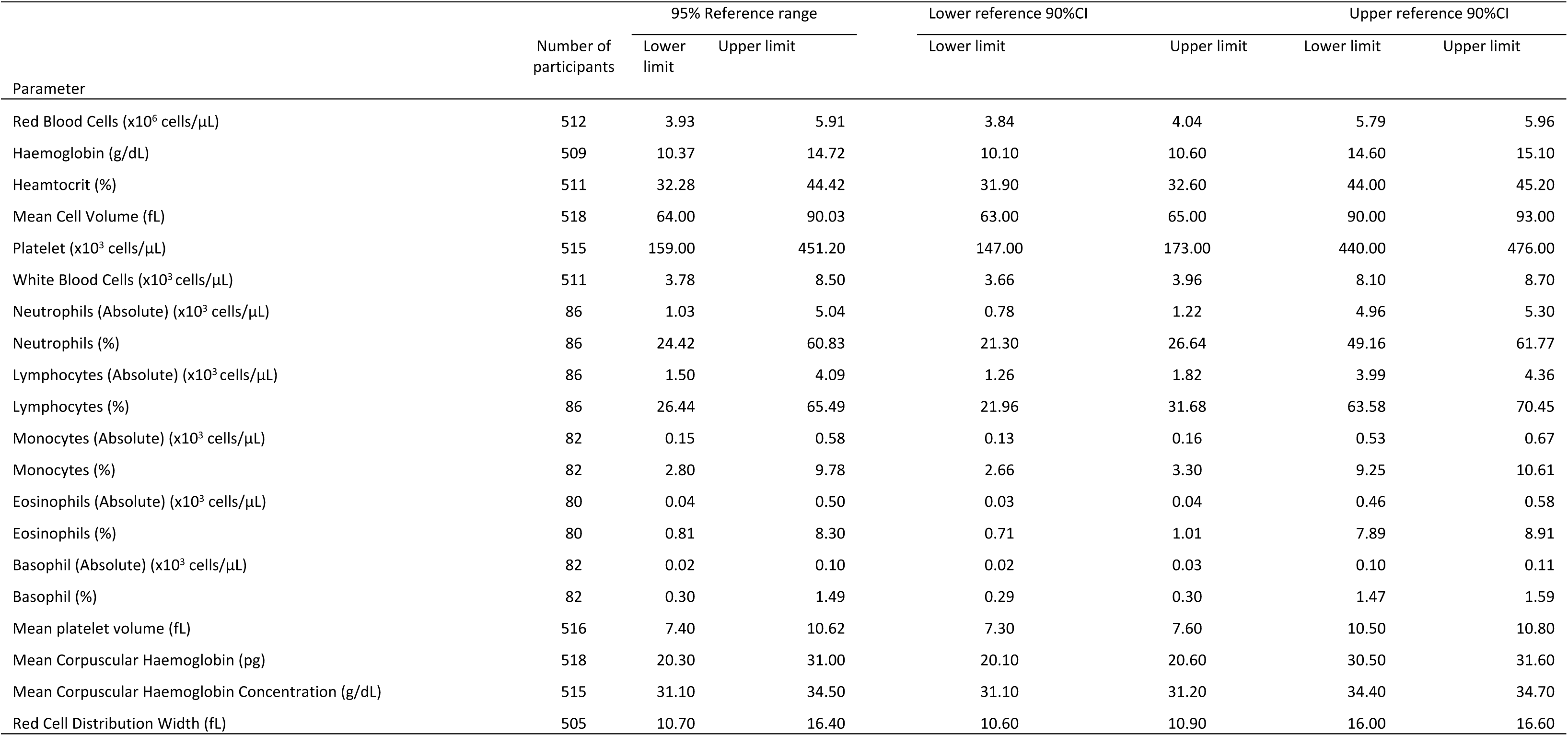
Reference ranges and associated 90% confidence intervals for **haematological** measures for healthy children aged **60-215 months** in Kilifi, Kenya, derived using data from clinical trials at KEMRI-Wellcome Trust Research Programme.

**Table 5a.**
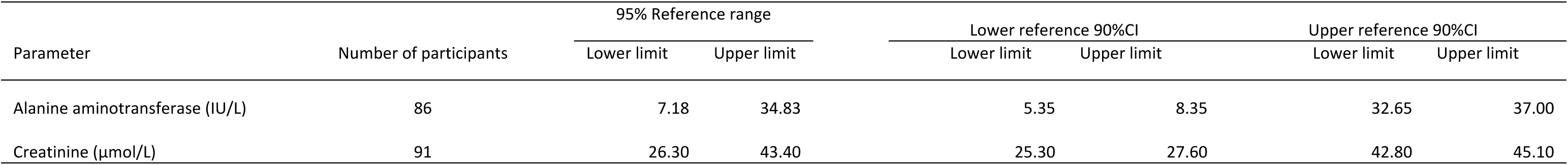
Reference ranges and associated 90% confidence intervals for selected **biochemical** measures for healthy **children aged 1-5 months** in Kilifi, Kenya, derived using data from clinical trials at KEMRI-Wellcome Trust Research Programme.

**Table 5b.**
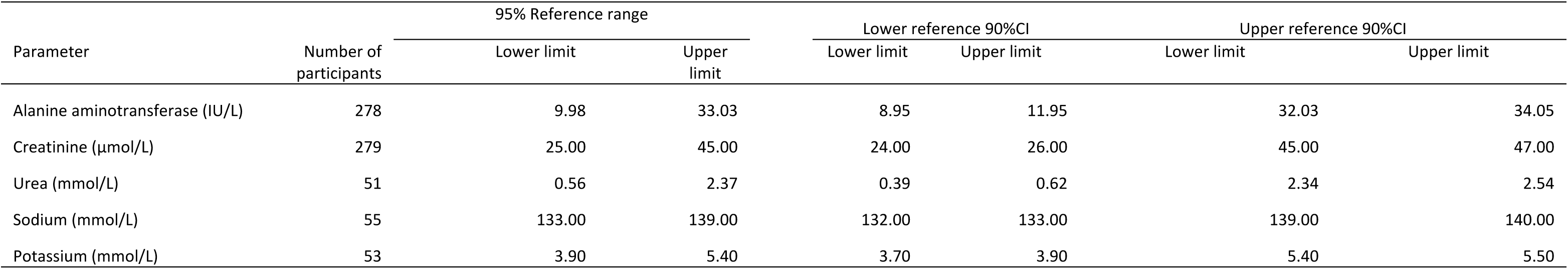
Reference ranges and associated 90% confidence intervals for selected **biochemical** measures for healthy **children aged 6-11 months** in Kilifi, Kenya, derived using data from clinical trials at KEMRI-Wellcome Trust Research Programme.

**Table 5c.**
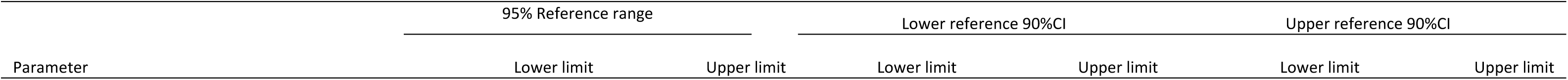
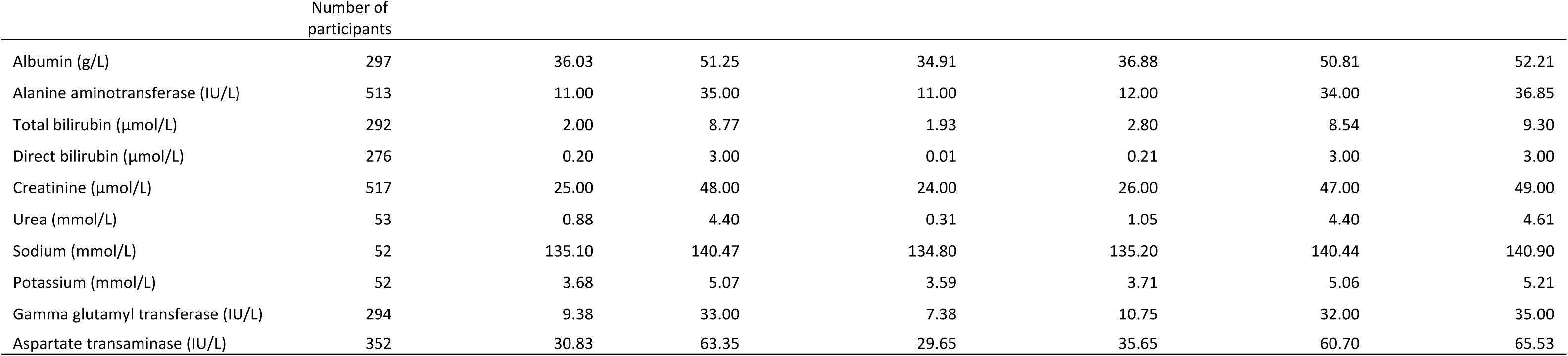
Reference ranges and associated 90% confidence intervals for selected **biochemical** measures for healthy **children aged 12-59 months** in Kilifi, Kenya, derived using data from clinical trials at KEMRI-Wellcome Trust Research Programme.

**Table 5d.** Reference ranges and associated 90% confidence intervals for selected **biochemical** measures for healthy **children aged 60-215 months** in Kilifi, Kenya, derived using data from clinical trials at KEMRI-Wellcome Trust Research Programme.

| Parameter | Number of participants | 95% Reference range |  | Lower reference 90%CI |  | Upper reference 90%CI |  |
| --- | --- | --- | --- | --- | --- | --- | --- |
|  |  | Lower limit | Upper limit | Lower limit | Upper limit | Lower limit | Upper limit |
| Urea (mmol/L) | 514 | 1.60 | 4.51 | 1.40 | 1.80 | 4.30 | 4.60 |
| Sodium (mmol/L) | 527 | 133.50 | 143.80 | 132.70 | 134.00 | 143.00 | 144.00 |
| Potassium (mmol/L) | 520 | 3.90 | 5.40 | 3.80 | 3.90 | 5.22 | 5.40 |
| Glucose (mmol/L) | 507 | 3.90 | 6.40 | 3.70 | 4.00 | 6.20 | 6.50 |
| Gamma glutamyl transferase (IU/L) | 328 | 12.00 | 37.55 | 11.00 | 12.00 | 35.00 | 38.00 |
| Phosphate (mmol/L) | 304 | 1.02 | 2.36 | 0.86 | 1.09 | 2.20 | 2.46 |
| Calcium (mmol/L) | 328 | 1.94 | 2.54 | 1.87 | 1.98 | 2.51 | 2.60 |
| Magnesium (mmol/L) | 271 | 0.72 | 1.31 | 0.69 | 0.75 | 1.25 | 1.35 |

### Comparisons with other locations

Comparisons between Kilifi 2023 adult and paediatric RRs with Kilifi 2017 RRs, other locations in Africa and elsewhere in the world are shown in **Fig 1 and 2, and Tables 6 and 7**. For some of these locations, parameters were not split by sex of participant, and some parameters were not available.

**Fig 2:**
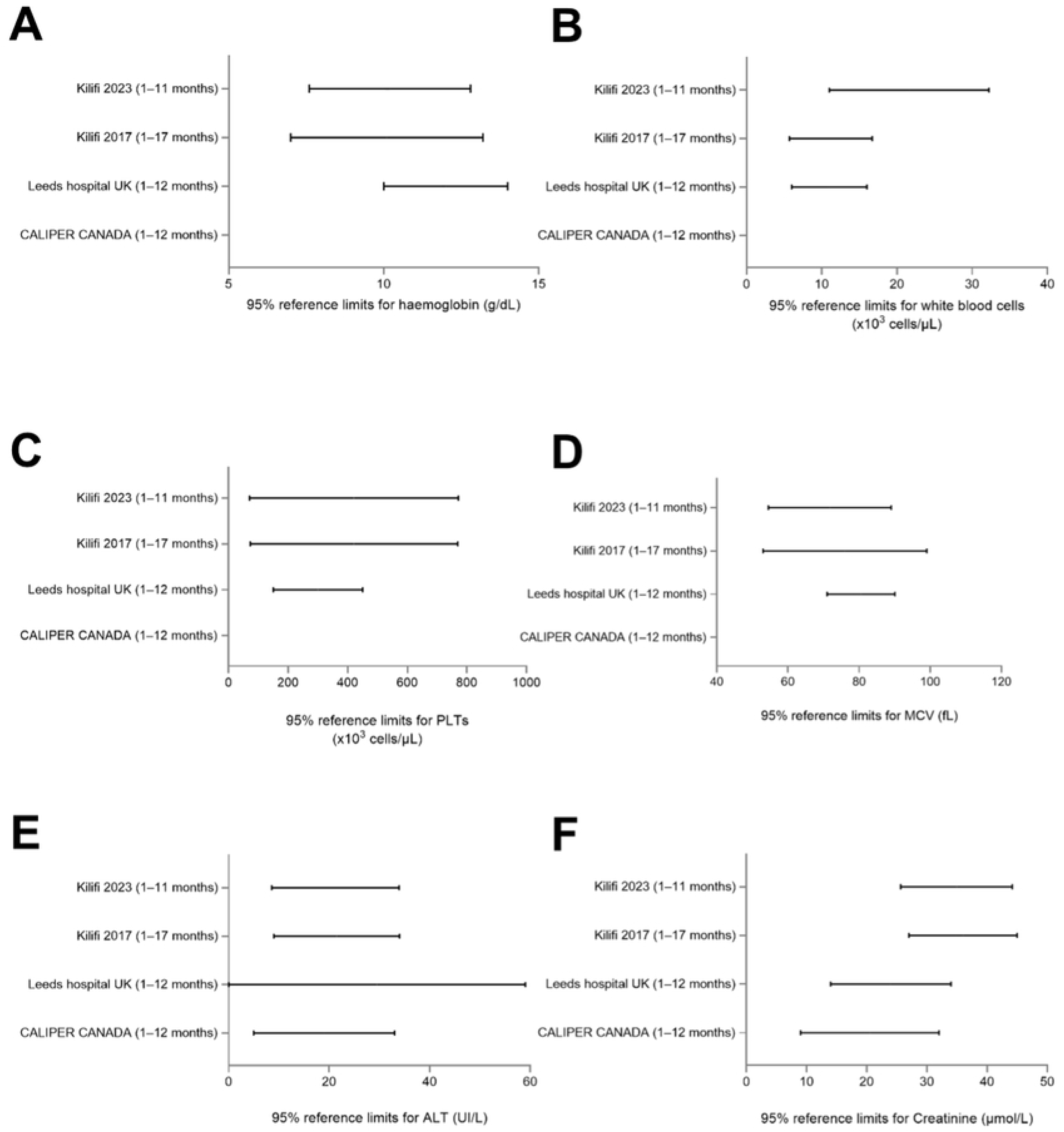
95% paediatric reference ranges of common parameters in 2023 Kilifi children aged 1–11 months compared with those from Kilifi 2017, United Kingdom and Canada: (A) haemoglobin, (B) white blood cells, (C) platelets (PLTs), (D) mean corpuscular volume (MCV), (E) alanine aminotransferase (ALT) and creatinine (F). UK = United Kingdom, CALIPER = Canadian Laboratory Initiative on Paediatric Reference Intervals.

**Table 6:**
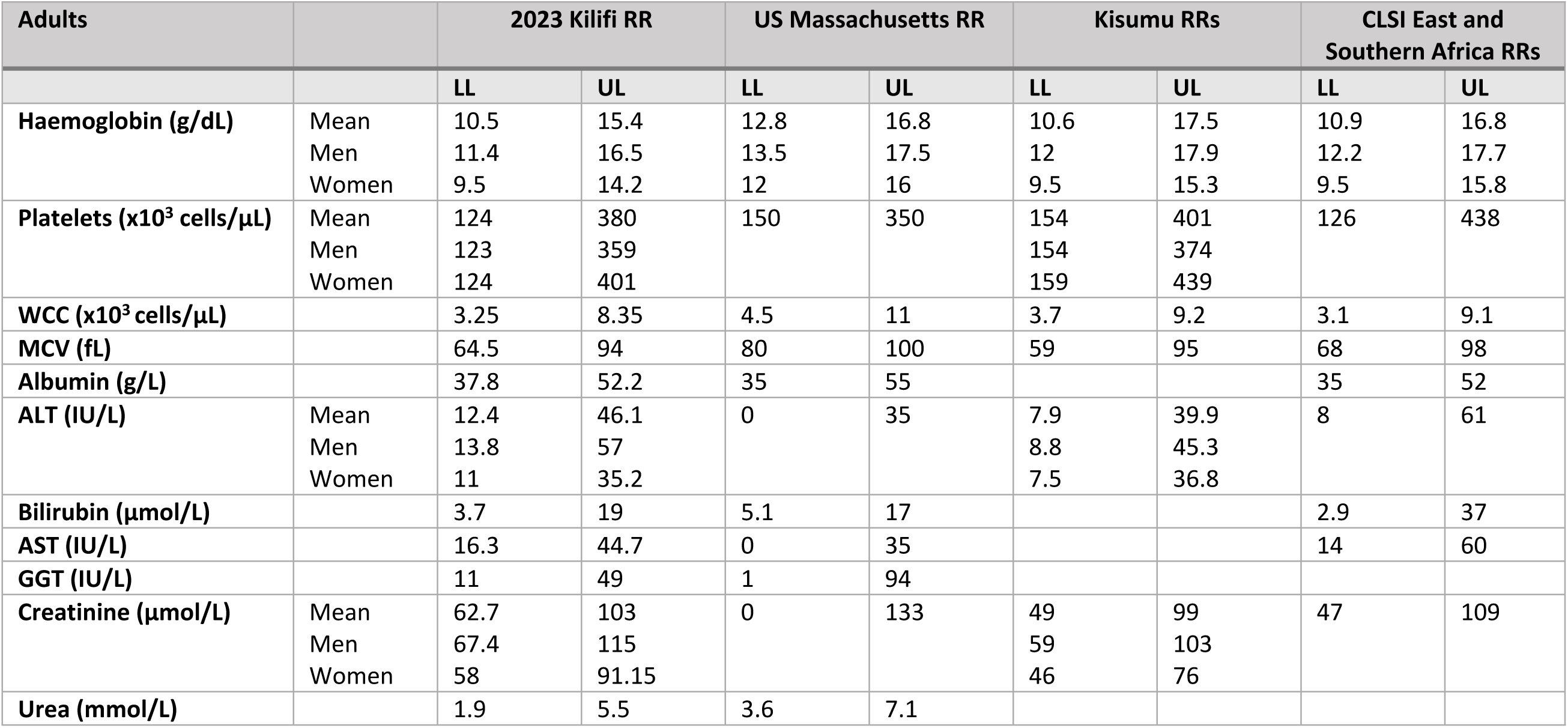
Comparing RRs for specific parameters from 2023 Kilifi ranges to those from elsewhere in Africa and the US. RR – Reference range; LL – Lower limit; UL – Upper limit; WCC – White blood cell count, MCV – mean corpuscular volume; ALT – Alanine aminotransferase; AST - Aspartate transaminase, GGT – gamma glutyltransferase,

**Table 7:** Comparison of current paediatric RRs both with previously derived Kilifi RRs from 2017 and with international RR’s. Most studies only derived RRs for children aged 1–12 months, so to enable comparison we averaged our 1–5 month and 6–11-month RRs to give the 1-12 month data. RR – Reference range; LL – Lower limit; UL – upper limit; UK - United Kingdom; WCC – White blood cell count; MCV – Mean corpuscular volume; ALT – Alanine aminotransferase.

| Children | 2023 Kilifi<br>1-12 months |  | 2017 Kilifi<br>1-17 months |  | Leeds Hospital UK<br>(1-12 months) |  | CALIPER Canada<br>1-12 months |  |
| --- | --- | --- | --- | --- | --- | --- | --- | --- |
|  | LL | UL | LL | UL | LL | UL | LL | UL |
| Haemoglobin (g/dL) | 7.6 | 12.8 | 7 | 13.2 | 10 | 14 |  |  |
| WCC (x10<sup>3</sup> cells/μL) | 5.5 | 16.1 | 5.7 | 16.7 | 6 | 16 |  |  |
| Platelets (x10<sup>3</sup> cells/μL) | 70.7 | 771 | 73 | 769 | 150 | 450 |  |  |
| MCV (fL) | 54.5 | 89 | 53 | 99 | 71 | 90 |  |  |
| ALT (IU/L) | 8.6 | 34 | 9 | 34 | 0 | 59 | 5 | 33 |
| Creatinine (μmol/L) | 26 | 44 | 27 | 45 | 14 | 34 | 9 | 32 |

### Adult populations

Adult 2023 Kilifi RRs were compared with those from Kisumu, Eastern and Southern Africa and the United States (17,18,20), Haemoglobin ULN in women was lowest in 2023 Kilifi RRs compared to other locations (14.2 g/dL vs 16 g/dL, 15.3 g/dL and 15.8 g/dL respectively). The overall ULN for ALT varied considerably being lowest in the US cohort (35 IU/L) and highest in the Eastern and Southern Africa cohort (61 IU/L). ALT ULN in 2023 Kilifi RRs was in between these two locations at 46 IU/L. Bilirubin was higher in the Eastern and Southern Africa cohort than in Kilifi (37μmol/L vs 19μmol/L respectively). ULN for creatinine was considerably higher in the US population at 133 compared to any of those from Africa.

### Paediatric populations

Paediatric 2023 Kilifi RRs were compared with those from Kilifi in 2017, United Kingdom and Canada (10,21,22). Firstly, when comparing our previously defined 2017 Kilifi RRs with 2023 Kilifi RRs, both were very similar. This is reassuring from a quality control perspective and adds weight to the accuracy of the current values. When looking further afield, the haemoglobin range was much wider in 2023 Kilifi RRs compared to the UK (ULN 12.8 g/dL and 14g/dL respectively. Platelet ranges were also much wider in both 2017 and 2023 Kilifi paediatric RRs compared to that of the UK (ULN 769× 10^3^/μL and 771 × 10^3^/μL compared with 450 × 10^3^/μL in the UK).

## Discussion

Here we have derived RRs for both adults and children from Kilifi County, Kenya to allow accurate interpretation of blood results for those participating in clinical trials at KWTRP. Previous studies have demonstrated the need for locally applicable ranges to avoid unnecessary exclusion from clinical trials, and to ensure blood abnormalities are correctly identified (4,5)

The number of parameters included in our RRs are more extensive than for other RRs either in Africa or more widely, and the sub-division of our paediatric RRs into different age groups is not represented elsewhere. Many other RRs use convenience or snowball sampling for participant selection, whereas here, random population sampling reduces the chance of sample bias. The differences observed here between men and women, along with between populations both within the African continent and further afield, support the development of sex and location specific RRs.

### Comparison of adult reference ranges

The differences we observed in Hb between men and women has been well described previously in many populations and has been proposed to be due to different hormone exposures in men and women, leading to improved oxygen delivery per unit of red cell mass in women, in combination with increased iron deficiency anaemia in menstruating women (23,24). The platelet differences observed here between sexes have also been well described previously although reasons for this are not well understood (6,25,26).

Increased liver function tests in men compared with women also fits with previous data but mechanisms behind these differences are complex and multifactorial (27). Environmentally, men tend to have an increased alcohol consumption compared to women predisposing them to alcoholic liver disease (28), and tend to have higher muscle mass than women contributing to elevated ALT and AST during muscle breakdown (29). Hormonally, oestrogens have a protective effect on the liver through various pathways. They influence glucose metabolism, alter fat deposition and lipid homeostasis in the liver, reducing the development of metabolic syndrome and non-alcoholic fatty liver disease (30,31).

When comparing our RRs with others in Africa, the much lower ULN of Hb in our adult population compared with that of Kisumu is particularly striking, despite both cohorts having a similar sex distribution. This is possibly due to the higher altitude in Kisumu compared to Kilifi (1131m vs 5m above sea level respectively) leading to higher levels of Hb in Kisumu for better oxygen transport when atmospheric oxygen is lower (32). Dietary patterns are also likely to vary between Kilifi and Kisumu, with Kisumu having the climate to grow a more diverse range of foods and having easy access to fish from Lake Victoria (33). Furthermore, there are likely socio-economic differences that may improve Hb in the population sampling in Kisumu. Both regions are endemic for malaria, which would reduce Hb levels, and Kisumu more endemic than Kilifi. However, there is marked heterogeneity of exposure in both locations, and the balance of urban vs rural populations including will impact the ranges of Hb seen.

The difference observed in adult liver function RRs between all African populations and the US population have been reported previously (4,17,34). Reasons for this are not clear, but suggestions include increased exposure to aflatoxin in Africa through poorly stored maize leading to Aspergillus growth (35,36), and variation in the pattern and type of alcohol consumption (37,38). This variability between populations indicates the importance of having a broad range of RRs to compare with other locations to identify discrepancies such as this (7).

### Comparison of paediatric references ranges

Differences between different age groups of children are as expected and as seen in other studies, including the variation in WCC and platelets with age (39). Other parameters remain relatively stable over time.

When looking at the variability between Kilifi and other locations, Hb is particularly lower in Kilifi children than those in the UK, however similar to that in other Africa studies. Reasons for this are multifactorial, but likely due to a combination of a higher incidence of malnutrition in Kilifi compared with the UK, along with increased incidence of chronic disease such as sickle cell anaemia and infections such as intestinal parasites or malaria (8). Platelet count RRs are considerably higher in all African RRs compared to those from the RCPCH UK. This may be due to chronic exposure to *Plasmodium* parasites in much of Africa leading to continued platelet activation (40), although recent acute malaria may also be associated with reduced platelet counts, and furthermore studies have shown that increased baseline platelet counts are protective against severe malaria through reducing parasite counts (41).

### Limitations

There are some limitations to how we have derived reference ranges, but generally these will be common to all methods of reference range determination and not unique here. Participants of these clinical trials are not fully screened for all diseases, only tested for infectious diseases and then clinically examined. There is no routine assessment for other issues such as undiagnosed cardiac disease or liver disease. Although participants here were recruited from the general population, these studies tended to recruit from a particular geographical area or specific population group so the selection will not be entirely random. Those volunteering for research studies are also likely to be more educated and engaged in clinical research than those not coming forwards. Their organ function therefore may be better than much of the population. A large proportion of women of childbearing age are also likely to have been excluded from these RRs due to the strict exclusion criteria applied to this group when recruiting to clinical trials.

Some of the adult biochemical parameters and paediatric parameters examined here did not meet the minimum recommended sample size of 120 for reference range determination, however many of these are not represented elsewhere in the literature, we feel they still hold value for presentation.

The last paediatric age group derived here includes a very large age range from 60–215 months. There may be some natural variation in blood parameters within this range, and this should be considered during interpretation.

## Conclusion

We describe the development of extensive adult and paediatric RRs in a coastal population in Kenya, demonstrating considerable regional variation both from other African RRs and RRs further afield. This indicates the need for locally derived RRs, and the minimal changes from our previously derived 2017 Kilifi paediatric RRs adds weight to the accuracy of these RRs.

## Data Availability

The data used in this manuscript is secondary analysis of existing data available from the respective clinical trials. Data is available either within the study cited papers, or will be available to researchers who submit requests to to gain access to the data following a signed data access agreement.

## Acknowledgements

We would like to thank all the principal investigators and sponsors of these studies for allowing reuse of their study data to generate these references ranges. Principal investigators: Prof Melissa Kapulu (CHMI), Prof Adrian Hill (Vac 073), Dr Mainga Hamaluba and Dr Josphat Kosgei (S4V01), Prof Anthony Etyang (ShinDa 2), Prof Anthony Scott (FPCV), Simon Kariuki (RTSS). Sponsors: University of Oxford (CHMI and VAC073), LimmaTech Biologics AG (S4V01), GlaxoSmithKline (RTSS), London School of Hygiene and Tropical Medicine (ShinDa 2 and FPCV). This manuscript was written with the permission of Director KEMRI CGMRC.

## References

1. Liu W, Bretz F, Cortina-Borja M. Reference range: Which statistical intervals to use? Stat Methods Med Res. 2021 Feb;30(2):523–34.

2. EP28A3C: Define and Verify Reference Intervals in Lab [Internet]. CLSI Standards. Available from: https://clsi.org/media/1421/ep28a3c_sample.pdf

3. O’Hara G, Mokaya J, Hau JP, Downs LO, McNaughton AL, Karabarinde A, et al. Liver function tests and fibrosis scores in a rural population in Africa: a cross-sectional study to estimate the burden of disease and associated risk factors. BMJ Open. 2020 Mar 31;10(3):e032890.

4. Price MA, Fast PE, Mshai M, Lambrick M, Machira YW, Gieber L, et al. Region-specific laboratory reference intervals are important: A systematic review of the data from Africa. PLOS Glob Public Health. 2022 Nov 14;2(11):e0000783.

5. Ichihara K, Ozarda Y, Barth JH, Klee G, Shimizu Y, Xia L, et al. A global multicenter study on reference values: 2. Exploration of sources of variation across the countries. Clin Chim Acta. 2017 Apr;467:83–97.

6. Biino G, Santimone I, Minelli C, Sorice R, Frongia B, Traglia M, et al. Age- and sex-related variations in platelet count in Italy: a proposal of reference ranges based on 40987 subjects’ data. PLoS One. 2013 Jan 31;8(1):e54289.

7. Achila OO, Semere P, Andemichael D, Gherezgihier H, Mehari S, Amanuel A, et al. Biochemistry reference intervals for healthy elderly population in Asmara, Eritrea. BMC Res Notes. 2017 Dec 19;10(1):748.

8. Quintó L, Aponte JJ, Sacarlal J, Espasa M, Aide P, Mandomando I, et al. Haematological and biochemical indices in young African children: in search of reference intervals. Trop Med Int Health. 2006 Nov;11(11):1741–8.

9. Number of trial registrations by location, disease, phase of development, age and sex of trial participants (1999-2022) [Internet]. [cited 2024 Mar 19]. Available from: https://www.who.int/observatories/global-observatory-on-health-research-and-development/monitoring/number-of-trial-registrations-by-year-location-disease-and-phase-of-development

10. Gitaka J, Ogwang C, Ngari M, Akoo P, Olotu A, Kerubo C, et al. Clinical laboratory reference values amongst children aged 4 weeks to 17 months in Kilifi, Kenya: A cross sectional observational study. PLoS One. 2017 May 11;12(5):e0177382.

11. Kapulu MC, Kimani D, Njuguna P, Hamaluba M, Otieno E, Kimathi R, et al. Controlled human malaria infection (CHMI) outcomes in Kenyan adults is associated with prior history of malaria exposure and anti-schizont antibody response. BMC Infect Dis. 2022 Jan 24;22(1):86.

12. Pharmacy and Poisons Board: Applications [Internet]. [cited 2024 Mar 19]. Available from: https://ctr.pharmacyboardkenya.org/applications/view/773

13. Etyang AO. Determining the Causal Role of Malaria in Elevating Blood Pressure and Pulse Wave Velocity in Kenyan Adolescents and Adults [Internet] [doctoral]. London School of Hygiene & Tropical Medicine; 2018 [cited 2024 Mar 19]. Available from: https://researchonline.lshtm.ac.uk/id/eprint/4646135/

14. Sang S, Datoo MS, Otieno E, Muiruri C, Bellamy D, Gathuri E, et al. Safety and immunogenicity of varied doses of R21/Matrix-M^TM^ vaccine at three years follow-up: A phase 1b age de-escalation, dose-escalation trial in adults, children, and infants in Kilifi-Kenya. Wellcome Open Res. 2023 Oct 12;8:450.

15. Efficacy of GSK Biologicals’ Candidate Malaria Vaccine 257049 Against Malaria Disease in Infants and Children in Africa [Internet]. [cited 2024 Mar 19]. Available from: https://clinicaltrials.gov/study/NCT00866619

16. Gallagher KE, Lucinde R, Bottomley C, Kaniu M, Suaad B, Mutahi M, et al. Fractional doses of pneumococcal conjugate vaccine - A noninferiority trial. N Engl J Med [Internet]. 2024 Sep 26 [cited 2024 Oct 1]; Available from: https://pubmed.ncbi.nlm.nih.gov/39330966/

17. Sing’oei V, Ochola J, Owuoth J, Otieno J, Rono E, Andagalu B, et al. Clinical laboratory reference values in adults in Kisumu County, Western Kenya; hematology, chemistry and CD4. PLoS One. 2021 Mar 30;16(3):e0249259.

18. Karita E, Ketter N, Price MA, Kayitenkore K, Kaleebu P, Nanvubya A, et al. CLSI-derived hematology and biochemistry reference intervals for healthy adults in eastern and southern Africa. PLoS One. 2009 Feb 6;4(2):e4401.

19. Scott JAG, Bauni E, Moisi JC, Ojal J, Gatakaa H, Nyundo C, et al. Profile: The Kilifi Health and Demographic Surveillance System (KHDSS). Int J Epidemiol. 2012 Jun;41(3):650–7.

20. Kratz Alexander, Ferraro Maryjane, Sluss Patrick M., Lewandrowski Kent B. Normal Reference Laboratory Values. N Engl J Med. 351(15):1548–63.

21. Adeli K, Raizman JE, Chen Y, Higgins V, Nieuwesteeg M, Abdelhaleem M, et al. Complex biological profile of hematologic markers across pediatric, adult, and geriatric ages: establishment of robust pediatric and adult reference intervals on the basis of the Canadian Health Measures Survey. Clin Chem. 2015 Aug;61(8):1075–86.

22. Leeds Teaching Hospital. Uk Leeds Blood-Sciences-Reference-Range-Database_BSHC-REC-317-v3.pdf.

23. Rushton DH, Dover R, Sainsbury AW, Norris MJ, Gilkes JJ, Ramsay ID. Why should women have lower reference limits for haemoglobin and ferritin concentrations than men? BMJ. 2001 Jun 2;322(7298):1355–7.

24. Murphy WG. The sex difference in haemoglobin levels in adults - mechanisms, causes, and consequences. Blood Rev. 2014 Mar;28(2):41–7.

25. Ranucci M, Aloisio T, Di Dedda U, Menicanti L, de Vincentiis C, Baryshnikova E, et al. Gender-based differences in platelet function and platelet reactivity to P2Y12 inhibitors. PLoS One. 2019 Nov 27;14(11):e0225771.

26. Patti G, De Caterina R, Abbate R, Andreotti F, Biasucci LM, Calabrò P, et al. Platelet function and long-term antiplatelet therapy in women: is there a gender-specificity? A “state-of-the-art” paper. Eur Heart J. 2014 Sep 1;35(33):2213–23b.

27. Ceriotti F, Henny J, Queraltó J, Ziyu S, Özarda Y, Chen B, et al. Common reference intervals for aspartate aminotransferase (AST), alanine aminotransferase (ALT) and γ-glutamyl transferase (GGT) in serum: results from an IFCC multicenter study. Clin Chem Lab Med. 2010 Nov;48(11):1593–601.

28. Vera MA, Koch CD, Liapakis A, Lim JK, El-Khoury JM. The ALT upper reference interval debate: Blame it on the alcohol. Clin Chim Acta. 2022 Feb 1;526:62–5.

29. Ma W, Hu W, Liu Y, He L. Association between ALT/AST and muscle mass in patients with type 2 diabetes mellitus. Mediators Inflamm. 2022 Oct 14;2022:9480228.

30. Kasarinaite A, Sinton M, Saunders PTK, Hay DC. The influence of sex hormones in liver function and disease. Cells [Internet]. 2023 Jun 11;12(12). Available from: 10.3390/cells12121604

31. Qiu J, Kuang M, He S, Yu C, Wang C, Huang X, et al. Gender perspective on the association between liver enzyme markers and non-alcoholic fatty liver disease: insights from the general population. Front Endocrinol. 2023 Dec 6;14:1302322.

32. Windsor JS, Rodway GW. Heights and haematology: the story of haemoglobin at altitude. Postgrad Med J. 2007 Mar;83(977):148–51.

33. FAO fisheries & aquaculture [Internet]. [cited 2024 May 23]. Available from: https://www.fao.org/fishery/en/news/41389

34. Kolahdoozan S, Mirminachi B, Sepanlou SG, Malekzadeh R, Merat S, Poustchi H. Upper Normal Limits of Serum Alanine Aminotransferase in Healthy Population: A Systematic Review. Middle East J Dig Dis. 2020 Jul;12(3):194–205.

35. Mekuria A, Xia L, Ahmed TA, Bishaw S, Teklemariam Z, Nedi T, et al. Contribution of Aflatoxin B1 Exposure to Liver Cirrhosis in Eastern Ethiopia: A Case-Control Study. Int J Gen Med. 2023 Aug 16;16:3543–53.

36. Afum C, Cudjoe L, Hills J, Hunt R, Padilla LA, Elmore S, et al. Association between Aflatoxin M₁ and Liver Disease in HBV/HCV Infected Persons in Ghana. Int J Environ Res Public Health. 2016 Mar 29;13(4):377.

37. Spearman CW. The burden of chronic liver disease in west Africa: a time for action. Lancet Glob Health. 2023 Sep;11(9):e1319–20.

38. Devarbhavi H, Asrani SK, Arab JP, Nartey YA, Pose E, Kamath PS. Global burden of liver disease: 2023 update. J Hepatol. 2023 Aug;79(2):516–37.

39. Li K, Peng Y-G, Yan R-H, Song W-Q, Peng X-X, Ni X. Age-dependent changes of total and differential white blood cell counts in children. Chin Med J (Engl). 2020 Aug 20;133(16):1900–7.

40. Watson JA, Uyoga S, Wanjiku P, Makale J, Nyutu GM, Mturi N, et al. Improving the diagnosis of severe malaria in African children using platelet counts and plasma PfHRP2 concentrations. Sci Transl Med. 2022 Jul 20;14(654):eabn5040.

41. Attaher O, Swihart B, Dang L, Santara G, Mahamar A, Keita S, et al. Higher platelet counts and platelet factors are associated with a reduction in Plasmodium falciparum parasite density in young Malian children. Int J Infect Dis. 2024 Feb;139:171–5.

42. Ngoto O. Data for: A Phase 1b, open-label, age de-escalation, dose-escalation study to evaluate the safety and immunogenicity of different doses of a candidate malaria vaccine; adjuvanted R21(R21/MM) in adults, young children and infants in Kilifi, Kenya [Internet]. Harvard Dataverse; 2023 [cited 2024 Mar 19]. Available from: 10.7910/DVN/IRGZ35

43. The Effect of Fractional Doses of Pneumococcal Conjugate Vaccines on Immunogenicity and Carriage in Kenyan Infants - Full Text View - Clinicaltrials.gov [Internet]. [cited 2024 Mar 19]. Available from: https://classic.clinicaltrials.gov/ct2/show/NCT03489018.

